# Growth Functions as a tool to model SARS-CoV-2 pandemic trajectory and related-deaths worldwide

**DOI:** 10.1101/2021.04.01.21254495

**Authors:** Vagner Fonseca, Edson Mascarenhas, Paulo Ramos, Leandro Coelho, Diego Frias

## Abstract

The international scientific community from different areas of knowledge has made efforts to provide information and methods that contribute to the adoption of the most appropriate measures to curb the spread of the COVID-19 disease. In particular, the data analysis community has been very active in publishing a large number of papers. A good part of them is related to the prediction of epidemic variables (number of cases and deaths) in different time horizons. To solve the problem of the prediction of COVID-19, an important place is occupied by the sigmoidal growth functions, as they have often been used successfully in previous epidemic outbreaks. The objective of this work was to investigate, on a statistical basis, the ability of classical growth functions to model the data from the COVID-19 pandemic. But for that, it was necessary to establish a clear classification of the 5 types of problems that can be faced with data analysis techniques in this specific context and to define a methodology based on quantitative metrics to measure the performance in solving these different types of problems. The basic concept used was that of an epidemic wave consisting of an initial-increasing and a final-decreasing phase. A classification of the COVID-19 waves in 4 types was done based on mining data from all available countries. Thus, it was possible to determine the resolvability of each type of problem depending on the stage of the epidemic wave. The biggest conclusion was the impossibility of solving the long-term forecasting problems (problem 5 – to estimate the total value of an epidemic wave) with data from the first phase only. Using this theoretical-methodological framework, we evaluated, using metrics specifically designed for these types of problems, the performance of 3 classic growth functions: Logistics, Gompertz and Richards (a generalization of the previous two) in 2 types of problems: (1) Description of the trajectory of the epidemic and (2) Prediction of the total numbers of cases and deaths. We used data from 10 countries, 7 of them with more than 100 daily deaths on the peak day. The results show a generalized underperformance of the logistic function in all aspects and place the Gompertz function as the best cost-benefit alternative, as it has performance comparable to the Richards function, but it has one less parameter to be adjusted, in the process of regression of the model to the observed data.

## Introduction

Severe acute respiratory syndrome coronavirus 2 (SARS-CoV-2) that causes the coronavirus disease 2019 (COVID-19), the respiratory illness responsible for the ongoing pandemic, has spread rapidly worldwide (Sohrabi *et al*., 2020; WHO, 2020a, 2020b). In October 30, 2020 the number of confirmed cases worldwide was approximately 45 million with 1.18 million deaths. After the peak in mid-April with an average of 7,500 daily deaths, the rate decreased to 4,300 daily deaths on average in late May, early June, increasing again until stabilizing in mid-June at 5,000 daily deaths, a rate maintained until the end October, the final date for updating this article.

Besides the proven virulence of SARS-CoV-2 there are some other factors that contribute to faster spreading:

a. Being an emerging pathogen, there is no pre-existing immunity in human population,
b. The symptoms are non-specific at early stage (mild respiratory symptoms and fever, 5-6 days after infection on an average) (WHO, 2020a) which difficult early identification and isolation of infected individuals,
c. It was found that some infected individuals can become contagious before symptoms onset, that is, even in the incubation period that is on average 5-6 days (Chen *et al*., 2020; Holshue *et al*., 2020; WHO, 2020b), and
d. The delay in the development of rapid diagnostic kits and subsequently the insufficient quantity of them to meet a global pandemic demand, which made it difficult for health authorities to control the epidemic in the countries that were first affected, among other factors regarding the promptness and efficacy of the health systems in different countries.

However, the number of COVID-19 daily tests in the world increased continuously every month with an average rate of ∼1.000 test by million (Wu *et al*., 2020). Such gradual increase in the number of the daily test is an exogenous factor that contributes increasing the number of daily confirmed cases worldwide and most be taken into account when evaluating the real number of infected people and the mortality of the pandemic.

The mortality of the SARS-CoV-2 virus is associated to development of acute respiratory distress syndrome, septic shock, metabolic acidosis coagulation dysfunction and organ failure in high risk patients (Chen *et al*., 2020; Holshue *et al*., 2020; Wu *et al*., 2020). Most patients with symptomatic COVID-19 were 30 to 79 years of age and mortality increases with age (Chen *et al*., 2020; Holshue *et al*., 2020; WHO, 2020a, 2020b; Wu *et al*., 2020). People aged over 60 years and those with underlying conditions such as hypertension, diabetes, cardiovascular disease have been found with highest risk for severe disease and death (Wang *et al*., 2020). The ICU patient mortality probability varies from less than 10% to more than 40% in different countries obeying to a set of simultaneous conditions that is not completely understood yet, especially because it is not known how the different risk factors influence the severity of the disease (Wang *et al*., 2020).

At a population level, the case fatality rate, *CFR*, defined as the ratio between the daily deaths, *d*(*t*), at day *t* due to COVID-19 and the daily number of confirmed (detected) cases, *c*(*t*), varies with the epidemic time *t* (in days) and differs among countries. In figure 1 it can be observed the evolution of *CFR*(*t*) in 10 countries and in the whole world since March 28, 2020 up to October 25, 2020. The time until death of the first serious cases causes a time lag to the right of the death curve in relation to the confirmed case curve, which causes an initial underestimation of the CFR in all countries, which is corrected slowly as epidemic time goes on. It is important to highlight that value of CFR is influenced by the number of tests, since the more tests are performed the greater the number of cases detected in the same population in the same moment. Now, taking into account that the number of deaths is independent of the tests and that the number of daily tests increased continuously in that period, it is to be expected a decrease in the CFR over time as observed in figure 1. However, there are other factors that can also lead to a decrease in CFR, such as, for example, an increase in the proportion of young people infected in relation to the elderly (Jason Oke, Daniel Howdon, 2020), the beginning of new epidemic waves, the improvement of prevention and life support therapies and the improvement of the capacity of the system health care to deal with the pandemic, among others.

**Figure 1.**
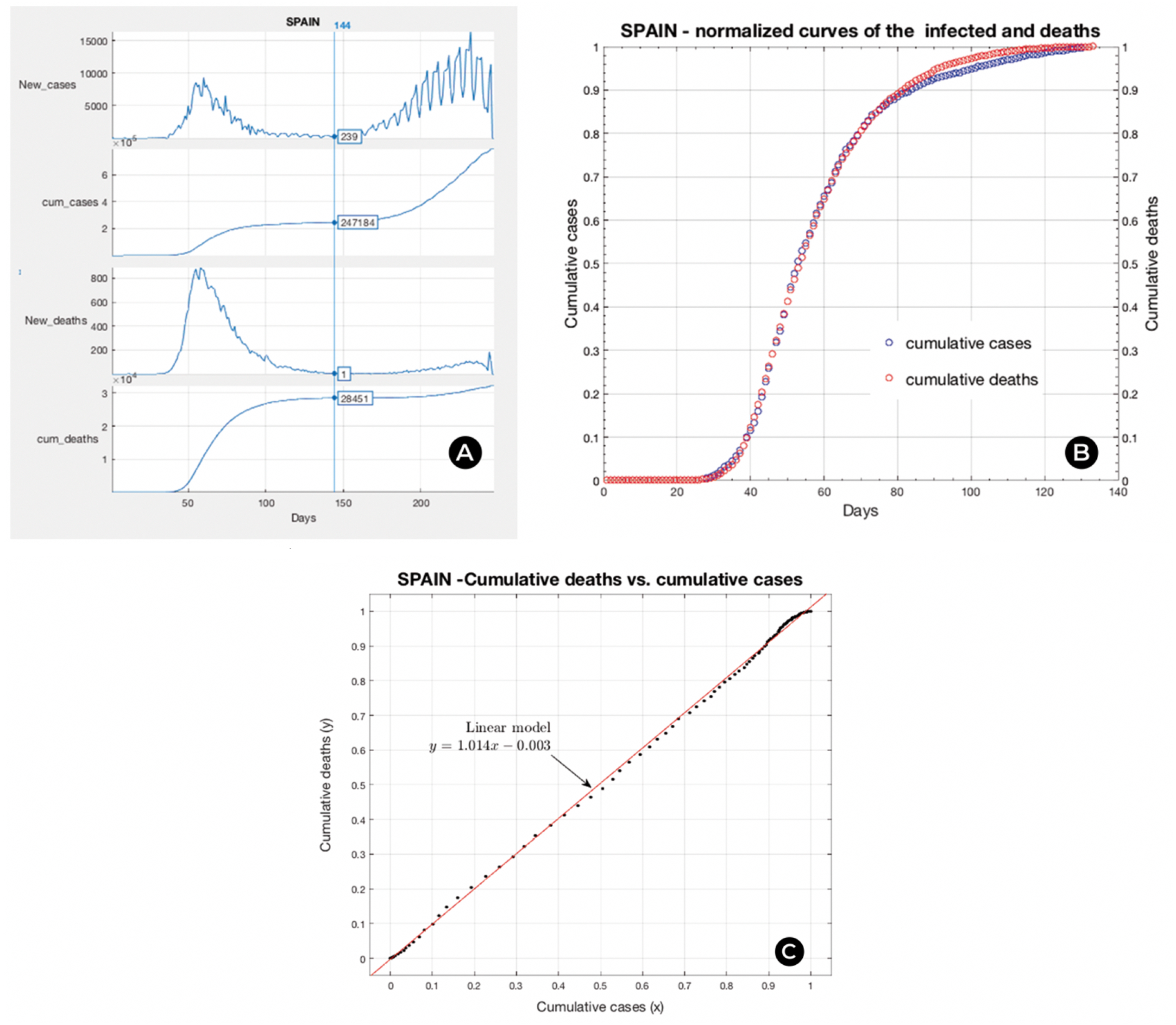
Case fatality rate of the ongoing COVID-19 pandemic. (Source: OurWorldInData.org/Coronavirus)

The COVID-19 pandemic has demonstrated once again that we need science in various academic disciplines in different combinations, to tackle the disease by limiting its spread and mitigating its consequences in the shortest possible time. In particular, information technology combined with the science of data analysis and modeling needs to answer key questions (limiting the analysis to 2 main variables: number of confirmed cases and deaths) that include: a) What is the current status? How many people are infected? How many are dying? How is evolving the epidemic? Where? and, b) What is the most likely situation in the future? How many more people will be infected? How many more will die? Where?

In the initial stage of an epidemic wave, there is an accelerated growth in the number of cases and daily deaths, erroneously called exponential, which is followed by a phase of decrease, forming a peak in the curves of these two variables. At this stage a more specific question needs to be answered: What will be the peak days and how many new cases and daily deaths will there be on those days? The correct answer to this question has transcendental relevance for the early preparation of the health system. From a logistical point of view, the public health system and society as a whole must be prepared for the number of clinical consultations, hospitalizations, autopsies and burials inferred from the data predicted for these critical days.

There are also other relevant questions that need to be answered, which are related to the total impact of an epidemic episode, in a country, in a region and worldwide. More specifically, in the case of a disease as lethal as COVID-19, it is important to predict the total number of deaths (death toll) caused by the pandemic. However, analyzing the pattern of both the daily number of new confirmed cases and deaths in all countries, we find a list of facts that must be taken into account before starting to model data for forecasting purposes. Even if most of these facts are known or intuitive, it is worth writing them in an organized way, as follows:

1. The temporal variation of epidemic variables describes waves,
2. An epidemic outbreak can produce several waves
3. In the case of multiple waves these can be superimposed or separated in time
4. A wave has two stages: the initial - increasing and the final - decreasing
5. The rate of growth / decrease can be classified as high or low. This classification needs global reference values, which can be obtained by working with data normalized to the maximum value in each country.
6. The data confirm that there are 4 types of waves, classifying them according to the intensity (low/high) of the rising - falling phases: symmetric patterns: low-low (Afghanistan) and high-high (Thailand), asymmetric: low-high (Egypt) and high-low (Italy), as shown in figure 2.

**Figure 2.**
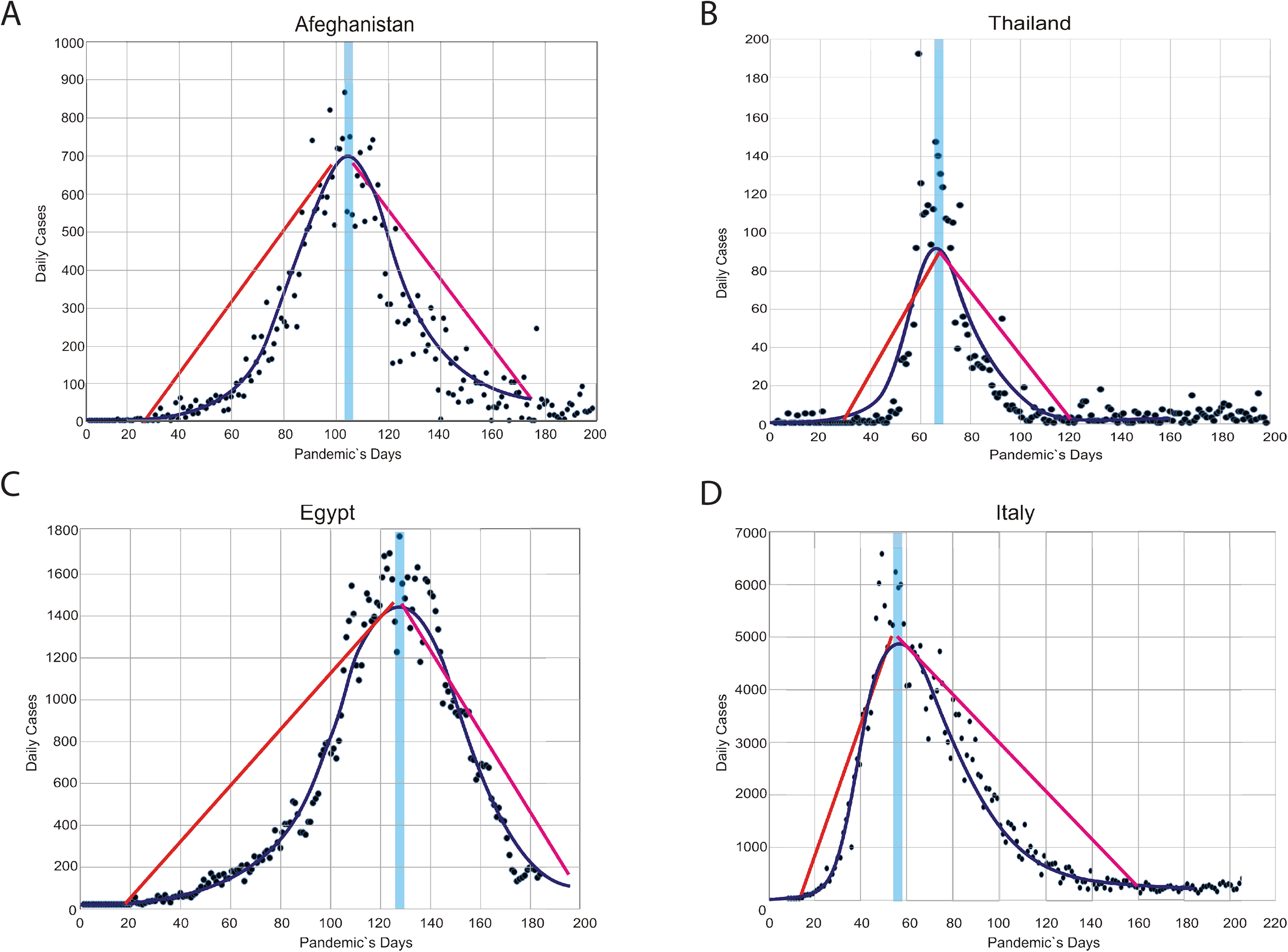
Basic Wave patterns: Left side: **Symmetric** slow-slow at top and high-high at the bottom. Right side: **Asymmetric** slow-high at the top and high-slow at the bottom.

These patterns found in data suggest independence between the two phases of the epidemic waves which suggest a hypothesis:

### Hypothesis

It is not theoretically possible to predict in a reliable way the shape of the second phase of the wave being in the first phase.

### Lemma

As a consequence, it is not possible to predict the total number of cases/deaths in the epidemic, with data only from the first phase, as it is necessary for that task to know the form of the second phase, which cannot be guessed in the first stage.

Based on this, we differentiate five categories of viable data analysis problems, described below:

1. **Epidemic trajectory description**: Real-time reconstruction, at any stage of the wave, of the epidemic trajectory using smooth curves that filter data fluctuation using some sliding window-based filtering or model-based regression technique. Public data shows that there are many countries with highly noisy data (highly fluctuating overnight) and downward and upward outliers that can make it difficult to recognize the current trend. It should be realized that the calculated curves are a hypothetic explanation of the underlying process.
  - **Suggested metrics**: Performance metrics for this task can be regarded to: (a) Degree of conservation in real time of the cumulative value of the described noisy variable, and (b) smoothness, which can be measured as being inversely proportional to the difference between the number of local extremes in the descriptive curve and observed in the real data, in a given inspection time interval.
2. **Short term prediction**: Forecasting epidemic variable from next day until 1-2 weeks in any wave stage.
  - **Suggested metric**: The natural performance metric for this task is to measure how accurate are the predicted values, depending on the anticipation of the forecast?
3. **Peak prediction**: To forecast the peak day and/or peak (maximum expected) epidemic variable value, being in the first wave stage.
  - **Suggested metrics**: This task can have two types of performance metrics: (a) How far ahead can a good forecast be made, for both peak day and value, and (b) How accurate are the predicted values depending on the anticipation of the forecast?
4. **Peak detection**: Real-time recognition of when the wave reached its peak. The intrinsic noise of the data and outliers can make it difficult to identify the wave itself and, correspondingly, its peak.
  - **Suggested metric:** The performance metric for this task should measure the relative delay in detecting the peak, for example, it can be given by dividing the days after the peak necessary for it to be identified by the days elapsed until the peak.
5. **Long term prediction**: Being in the second phase of the wave (after the peak day) recognize the pattern of decrease to predict the total values of the epidemic variables in the wave under study.
  - **Suggested metrics**: This task can have two types of performance metrics: (a) How far ahead can a good forecast be made and (b) How accurate are the forecasted values?

Different types of models can be used for solving problems 1-3 and 5, as well as machine learning methods. The scope of our study is the investigation of the solution of the first (in the second wave stage) and fifth categories of problems using growth models. Population dynamics models are particularly useful for describing epidemic variables, i.e. the number of cases and deaths, because (Bürger, Chowell and Yissedt Lara-Díıaz, 2019):

1. They are simple ordinary differential equations (ODE) that can be solved explicitly under certain assumptions about the epidemic process that is being modeled, obtaining the so-called growth functions,
2. The description of the underlying biological and social interaction mechanisms can be avoided, leading to a small number of model parameters that can be easily estimated by regression against the collected data, and
3. Can be used for efficient and fast forecasts with quantifiable uncertainty, even with highly noisy data.

In this work we addressed the use of such kind of models, more specifically of the growth functions for both purposes: describing the previous and actual situation and predicting the future of the studied variables in corona virus epidemics. Our base model was the 4-parameters Richard’s growth function (Pienaar and Turnbull, 1973; Tsoularis, 2001) which becomes Logistic and Gompertz when one of its parameters approaches 1 and 0, respectively. Logistic and Gompertz growth functions have three parameters and described the cumulative epidemic variables.

Some basic knowledge of analytical geometry is necessary to read this article. The temporal derivatives of the growth function are the daily rate of the epidemic variable. Accordingly, there are two analogies that need to be taken into account: (1) The inflection point of the growth function that describes the path of the cumulative epidemic variable is the peak point of the corresponding daily rate curve for this variable, which has a waveform, and (2) The asymptote (maximum value) of the modeled cumulative variable is equal to the total number on the wave, which is mathematically equivalent to the time integral (area below the curve) of the daily curve (wave).

We use publicly available data from COVID-19 from 10 countries having different curve shapes and having passed the first wave (Xu *et al*., 2020). A novel method was implemented to estimate how many days before the practical end of the wave the different growth functions were able to predict the final number of deaths in each country within a preset error of +/-10%. The descriptive and predictive results obtained with each model were compared and the most robust function in each country was identified, as well as the best choice in the group of countries studied. For comparison of the long-term predictive performance (problem 5) a set of metrics was proposed.

In order to evaluate the agreement of our results with previous works, we conducted an extensive web search for peer-reviewed recent articles since 2015 using a very detailed and specific search string containing set of terms that determines the subject (coronavirus outbreaks), the model (growth functions) and the application (prediction of epidemic variables), adding a set of terms to exclude preprints and other sources of PDF files. Our search did not include books or theses.

This search returned 19.450 matches, 18.200 (93.5%) of which were published in 2020 related to the COVID-19 pandemic. After visual inspection and reading of abstracts, some articles were selected with better correspondence according to five categories: (1) Growth functions studied / applied, (2) Measured performance criteria, considering two classes: descriptive and predictive, (3) Epidemic variable modeled, considering two classes: confirmed cases and deaths, (4) outbreaks / viruses studied, and (5) objective of the study, considering two classes: comparative and declarative. This procedure led to a reduced set of 29 relevant and consistent articles, but only a few of them will be discussed in the next section due to space limitations and partial overlap of content.

It should be noted that in none of the related works found, was any reference to any type of classification or categorization of the problems addressed from the perspective of data analysis, nor the use of metrics equivalent to those described in this article.

## Related works

In (Verma *et al*., 2020) data on confirmed cases of COVID-19 in seven countries, viz., Germany, China, France, United Kingdom, Iran, Italy and Spain, and in the World as a whole, have been adequately described using a generalized linear procedure followed by regression with Gompertz function. They studied the impact of lethal duration of exposure on mortality rates. Unfortunately, the article is not conclusive and the correlation sought is not shown. According to our problem classification, they approached the type 1 problem to solve the type 5 problem for a derived variable: lethal duration of exposure. As they introduced a new derived variable, there are no previous reference values to analyze the performance of the method used. In addition, their results are based on assumed mortality rates equal to the Case Fatality Rates (CFR) data shown in figure 1, until March 21, 2020. As noted above, the CFR at that time was underestimated, stabilizing later, before a subsequent decay, which invalidates the approach used.

In (Dutra *et al*., 2020) the Gompertz function is used to describe the evolution of the total number of deaths by COVID-19 in Brazilian states (problem 1) and to predict the number of deaths (problem 5), which is the same predictive variable that we address. However, our study is more extensive considering three growth functions and 10 countries. They show that the forecast of the total number of deaths in a wave can only be made in states where the peak of the wave occurred (confirming our hypothesis), but they did not measure the suggested metrics as is done in this article. In addition, in states where the peak of the wave did not occur, they try to extend the method using a predicted peak day, whose accuracy could not be confirmed.

In (Kamrujjaman, Mahmud and Islam, 2020) the authors compared the Logistic and Malthusian (exponential) growth functions to model daily confirmed case data worldwide and in China. At the time of the analysis, the world was starting the first stage of the wave and China was reaching its peak. Therefore, the problem addressed is of type 1 in the first initial stage of the wave according to our classification. They found a better descriptive performance of the Logistics function than the Malthusian model, mainly in China. However, this result could have been easily inferred by analyzing the situation and the models chosen: The Malthus model is not able to describe waves as expected and was later confirmed in the pandemic COVID-19. In addition, no metrics were used showing only figures where the data and curves generated with the two models were plotted.

In (Dutra, Farias and Melo, 2020), a new approach is proposed to predict the total number of infected cases, that is, problem 5. In this article published in May / June 2020 (less than 4 months ago), we find the phrase “the models used in previous works (Dutra *et al*., 2020) they were good at adjusting the observed data, but their predictions have not been confirmed”. Translating to our knowledge structure, it is equivalent to saying “the previous models were good for problem 1, but not for problem 5”, which, correcting that the problem was not in the models but at the moment and objective for which they were applied, only confirms our hypothesis that no model is able to reliably solve problem 5 using only data from the first stage of the epidemic wave. Their method combines the functions of Logistic growth and Gompertz, which provide lower and upper limits, respectively, of the sought total number of infected cases in the epidemic. They show that the weighted average of such limits is a good predictor, reaching an accuracy> 85% in the eight countries studied. It is important to realize that the eight countries used as an example (Australia, Austria, China, Croatia, New Zealand, South Korea, Switzerland and Thailand) were at the time ending the second phase of the first epidemic wave, which is evidence of viability of problem 5 solution for waves in the second phase. In part, the choice of countries where the first wave was completed can be understood as necessary to have a data reference to calculate the forecast accuracy of the total number of cases in the first waves of those countries. However, in the real-time simulation the predictive method was applied also during the first wave stage in these countries leading to the following conclusion “it can be concluded that non-linear regression adjustments of both the Gompertz function and the Logistic function before the inflection point (our comment: that is, before the peak point of the wave of daily cases) do not result in a good determination of the maximum limit for the number of accumulated cases”. This empirical finding supports the above Lemma.

Although the cited diverging aspects, both works share the same method of real-time monitoring the growth function parameter that defines the plateau of the curve of the cumulative studied variable. Monitoring is done, in both cases, by a daily regression of the model to the available data. Therefore, from a methodological point of view (Dutra, Farias and Melo, 2020) is the most similar article found in our review process.

Finally, in (Bürger, Chowell and Yissedt Lara-Díıaz, 2019) the authors investigate the performance of four growth functions: Logistics, Gompertz and their generalized versions (replacing the number of cases by its 0,1 power), when modeling epidemic data. More specifically, they measured the descriptive capacity of each model and then they addressed the type 1 problem. They used 37 epidemic outbreak data sets, in order to identify the most appropriate growth model for each case to describe epidemiological data. They observed that the Generalized Logistics model outperformed the other models in describing the vast majority of epidemic trajectories.

Taking into account that they were particularly interested in obtaining information about the variant of the model that provides a better description of the different epidemic outbreaks, it was considered the most similar study regarding the scope of the work. However, to better elucidate the similarities and differences between them, in Table 1 we show a detailed comparison according to the five categories used to characterize the previous works.

**Table 1.** Detailed comparison of this work with the article (Bürger, Chowell and Yissedt Lara-Díıaz, 2019) found to be the most similar in scope.

| Category | Base article | This work |
| --- | --- | --- |
| Measured performance criteria | descriptive performance | descriptive and predictive performances |
| Studied growth functions | Logistic and Gompertz and respective generalizations | Richards with Logistic and Gompertz as particular cases |
| Data (epidemic variable) | Confirmed cases in 37 epidemic outbreaks | Deaths in 10 countries |
| Outbreaks / virus | HIV, Yellow fever, Malaria, others | COVID-19 |
| Common purpose (scope): Comparison of the performance of growth function on modeling different epidemic trajectories. |  |  |

According to the authors’ knowledge, this is the first time that Richard’s model, a generalization of the two most commonly used growth functions, Logistics and Gompertz, is extensively evaluated using a representative set of countries, both in terms of its accuracy in describing the trajectory of the epidemic, as well as in its ability to predict accurately and in advance the total number of deaths in the COVID-19 pandemic.

In addition, no classification was found related to data analysis problems in epidemic episodes or to the specific set of performance metrics introduced in this work.

Furthermore, no hypothesis was found in the recent bibliography (2015-2020) regarding the impossibility of solving long-term prediction problems with data only from the first phase of the epidemic wave, presented in this work.

## Materials and Methods

### Data Collection

Time series with daily data from the COVID-19 pandemic in all countries was downloaded from the public repository of the World Health Organization on 2020-08-30 (WHO, 2020c). In particular, we processed the series with the number of confirmed cases and deaths, as we intended to conduct research in both series to assess possible differences in results.

Taking into account that the multi-wave character of the pandemic had already manifested itself at the time of the survey, and that the growth functions describe the dynamics of a single wave, it was necessary to establish criteria to select countries and time periods, with a single wave complete.

For this, we established the criteria for the beginning and end of the wave and the minimum size of the wave to be chosen. The first day of the wave was the day of the first confirmed case or death, depending on the series and the last day was defined as the first day after the peak with a daily number of confirmed cases, respectively deaths, below a threshold set at 1% the peak size. Following these criteria, setting the minimum size of the first peak in the daily number of deaths at 100, seven countries were selected: China, Denmark, France, Germany, Spain, Sweden and the United Kingdom. Subsequently, in order to analyze the performance of the models in countries with peaks below 100 daily deaths, three countries were added to the list: Finland, Greece and New Zealand.

## Data Pre-analysis

Before conducting the modeling study with the growth functions of the cumulative confirmed cases and cumulative deaths curves, a comparative study of their forms was carried out in each country. The objective of this phase was to determine whether it was really necessary to model the two series or not. In other words, if there was a very high linear correlation between the two curves, then it would be pointless to model both, since the results would also be very similar.

Figure 3 shows the results of this comparison for the case of Spain. A very high linear correlation (Pearson-r> 99%) was observed between the normalized cumulative curves of confirmed cases and deaths. Normalization was done by dividing by their respective maximum values in the considered time interval. In part A, we show the daily (above) and cumulative (below) number of new confirmed cases (upper half) and deaths (lower half) for Spain. The vertical line on day 144 defines the end of the data set that contains the first wave in this country. In part B, we represent the overlap of the cumulative number of confirmed cases and deaths after normalization. In part C, we show the scatter plot of the normalized cumulative number of confirmed cases and deaths and linear regression, showing that the agreement between the normalized series is almost perfect, giving a Pearson-r very close to 1 (Table 2).

**Table 2.** Correlation between cumulative confirmed cases and cumulative deaths.

| Country | Pearson-r |
| --- | --- |
| China | 90,86% |
| Denmark | 99,65% |
| Finland | 98,54% |
| France | 99,03% |
| Germany | 95,52% |
| Greece | 98,87% |
| New Zealand | 80,37% |
| Spain | 99,98% |
| Sweden | 94,46% |
| United Kingdom | 99,60% |

**Figure 3.**
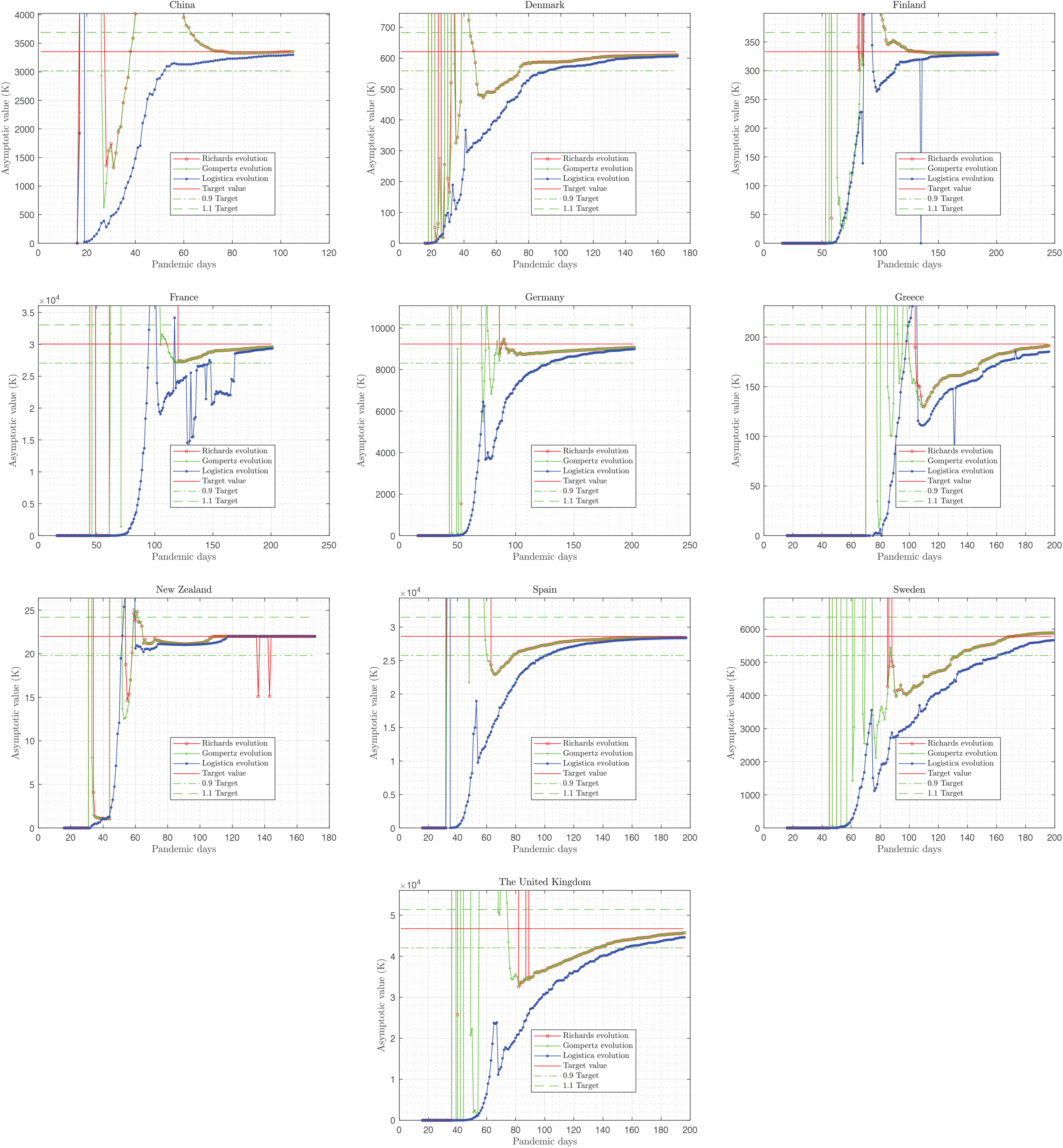
Epidemic curves of Spain: **A)** curves of confirmed cases (above) and deaths (below), both daily and cumulative. **B)** Normalized curves in the time interval from 12 to 144 days. **C)** Scatter plot of the cumulative confirmed cases vs. cumulative deaths for Spain.

The results were similar in all countries, regardless of the peak size. Table 2 shows the linear correlation rates between confirmed cases and cumulative deaths in all selected countries. According to these results, we decided to deal only with the time series of deaths in our study. At this point, it is important to note that when we refer to the epidemic data, process or its trajectory in a given country, we are actually referring to the daily or cumulative number of deaths in the first complete epidemic wave in that country.

## Considered Growth Models

Three growth models were investigated in this work, whose equations and characteristics are described below.

- **Logistic function** The logistic model generates a symmetrical sigmoid curve that describes the growth in the number of cumulative deaths over time. Let us denote by *t* the time (in days) and let *N(t)* be the number of deaths until day *t*. The logistic growth function is given by the equation:

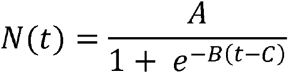

where parameter *A* is the final size of the epidemic (upper asymptote), *B* is the growth rate and *C* is the inflection day on the curve. When *t = C*, the number of deaths is 50% of the final number of deaths *A*, which reflects the symmetry of the logistic function.
- **Gompertz function** In contrast to the logistic model, the Gompertz function is asymmetric, showing greater growth in the vicinity of the lower asymptote, or small values of *N*. It is given by the equation:

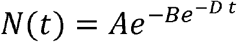

where parameter *A* is the final epidemic size (upper asymptote), and *B* and *D* are related to the rate of growth and inflection day of the curve, which in this case is given by

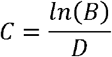 When *t = C* in this case, the number of deaths is *N(C)=A*/*e* which is approximately 36.8% of the final death toll *A*, reflecting the asymmetry of the Gompertz function.
- **Richard’s function** Richard’s function is a variant of the sigmoid function that generates more flexible S-shaped curves and was derived from the Von Bertalanffy growth function (Pienaar and Turnbull, 1973). Its four-parameter version is given by the equation:

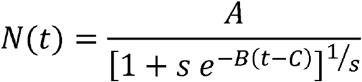

where the parameter *A* is the final epidemic size (upper asymptote),*B* is the growth rate, *C* is the inflection day and the parameter *s* varies in the interval ]0,1]. Note that when *s*=1 the Richard’s function becomes the Logistic function and when *s*→0^+ it becomes the Gompertz function.

### Epidemic Trajectory Classification

To identify which model best fits the epidemic data for each of the 10 countries studied, a series of regressions was performed with all available data (daily cumulative number of deaths), using the Richard’s model, with the parameter *s* varying from a value very close to zero (equal to the accuracy of the real machine) up to 1, with step = 0.01. The value of *s* that minimized the coefficient of determination R-squared was considered the best choice for each country. This way, when the best choice of *s* was close to zero, the Gompertz function was chosen as the best, while when the best *s* was 1, the logistic function. For 0 <*s* <1, Richard’s function was chosen. A code written in Matlab scripting language was used for this purpose.

### Dynamic Non-linear Regression

After the classification of the epidemic trajectory of each country, we performed a simulation of the respective epidemic trajectories, starting 10 days after the day of the first death and adding one more day until reaching the end of the data set in the country. Each day the model chosen with the optimal value of s is adjusted to the data in the corresponding time interval, calculating the other three parameters *A, B* and *C* (or *D*).

### Dynamic Description of the Epidemic Trajectory

Evaluating the model with the optimized parameters, the theoretical curve can be plotted daily with the observational data, allowing to visualize the descriptive capacity of the chosen model over time.

### Dynamic Prediction of the Death Toll

Parameter *A (t)* calculated each day *t*, is a forecast of the total number of deaths, *D*, at the end of the epidemic. For this reason, we denote *D*_*pred*_(*t*)=*A(t)* in the remaining text. It was observed in the simulations with real data (shown in the Results section) that at the beginning of the epidemic episode, the predicted value fluctuates from day to day until it starts to vary smoothly and converges monotonically to a value very close to the real number of deaths, *D*, in all cases.

To measure the predictive capacity of the studied models, we introduced a metric called days of anticipation, *d*_*ant*_, which is the number of days before the end of the wave in which the predicted number of deaths differs from the known value (in our experiments), in less than a given error tolerance *ε* (given as a percentage). Mathematically, it occurs on day *t*=*d*_*p*_, if

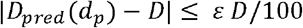

and then *d*_*ant*_= *d*_*end*_ *-d*_*p*_, where *d*_*end*_ is the final day of the wave. In this article we use *ε* = 10%.

However, a single but necessary condition must be satisfied to accept *d*_*ant*_ as the days of anticipation in an experiment. The condition guarantees that after the day considered the forecast is close enough to the real value, the forecast never gets worse until the end of the wave. Mathematically, it must be satisfied that | *D*_*pred*_(*t*) - *D*| ≤ *ε D*/100 for all days after *d*_*p*_, that is, for every *t > d*_*p*_.

In addition, taking into account that the waves have different durations, a relative measure is recommended to compare the predictive performance of each method in different waves. For example, 30 days in advance may be significant for an 80-day wave, but not as significant for a 200-day wave. For this, we introduce the relative anticipation (given in%), denoted *R*_*ant*_ by, defined as

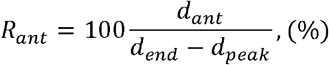

where is *d*_*peak*_ the day of the peak of the wave drawn by the daily deaths data.

As discussed earlier, we hypothesized that it makes no sense to try to predict the total number of deaths before the start of the second decreasing stage of the epidemic wave. As the second stage begins at the peak day, is always less than the length of the second wave stage given by *d*_*end*_ − *d*_*peak*_, that is, *d*_*ant*_*≤ d*_*end*_ − *d*_*peak*_. Under this assumption it holds that *R*_*ant*_ *∈* [0,100].

In order to measure the final accuracy of the forecast, the value predicted at the end of the simulation, *D*_*pred*_ *(d*_*end*_), is compared with the number of deaths accumulated on the last day of the time series, *D*. The final accuracy is defined as:

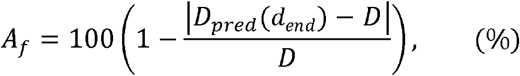

It should be noted that, satisfied the necessary condition stated above, the initial accuracy of the prediction *A*_*f*_(*d*_*p*_) is always less or equal the tolerance *ε* used. Therefore, it holds that *A*_*f*_(*t*) ≥ 100 *− ε* for all *t* ≤ *d*_*p*_.

## Results

The results of the study will be presented separately to differentiate those of the evaluation of the descriptive capacity from those of the predictive capacity. We understand as descriptive capacity the ability of the theoretical model (in our case the growth functions studied) to accurately describe the observed data, from the beginning of the process (isolated pandemic wave) until the date of observation. In particular, in the application being evaluated, we measure the quality of both the description of the number of cumulative deaths directly by the growth function under study, and the description of the daily number of deaths by the time derivative of this growth function. As the measure of the agreement between observational and theoretical data we use R-square. Thus, the higher R-square, the greater the descriptive capacity of the model.

On the other hand, the predictive capacity of the model is determined, in the first instance, by the similarity between the data to be observed in the future and the theoretical prediction, calculated with the adjusted analytical model (mathematical formula), for the respective days ahead.

Obviously, the predictive capacity should improve as more data (days) are available (considered) to adjust the model, which brings up a second predictive capacity criterion. This criterion is related to how many days before the end of the pandemic the model is able to make an accurate prediction, within a certain error limit, of the remaining trajectory of the epidemic process.

In this work, we chose as a target the total number of deaths, *D*, in an epidemic wave, in order to be able to compare the different growth models, both for the accuracy of the model in the prediction of *D*, and for the days in advance with which each model makes a consistent (which does not worsen afterwards) and accurate (within a certain error range - 10% in the present study) prediction of *D*.

The descriptive and predictive results are discussed in the next sections.

### Results of the Descriptive Capacity of the Growth Models

In this section, we present results that allow us to answer the questions: How well do the models studied describe the trajectory of the epidemic in the selected countries? What is the best model? What are the descriptive limitations of these models?

Table 3 lists the R-square values for the three models that describe the temporal evolution of the cumulative number of deaths considering the first complete epidemic wave in each country. It should be noted that all growth functions performed very well (R2> 91%) from a descriptive point of view. However, the Richards function has the best average R^2^ for the countries studied, being significantly better in Germany and the United Kingdom, where the other two functions performed less than 93% and 92%, respectively.

**Table 3:**
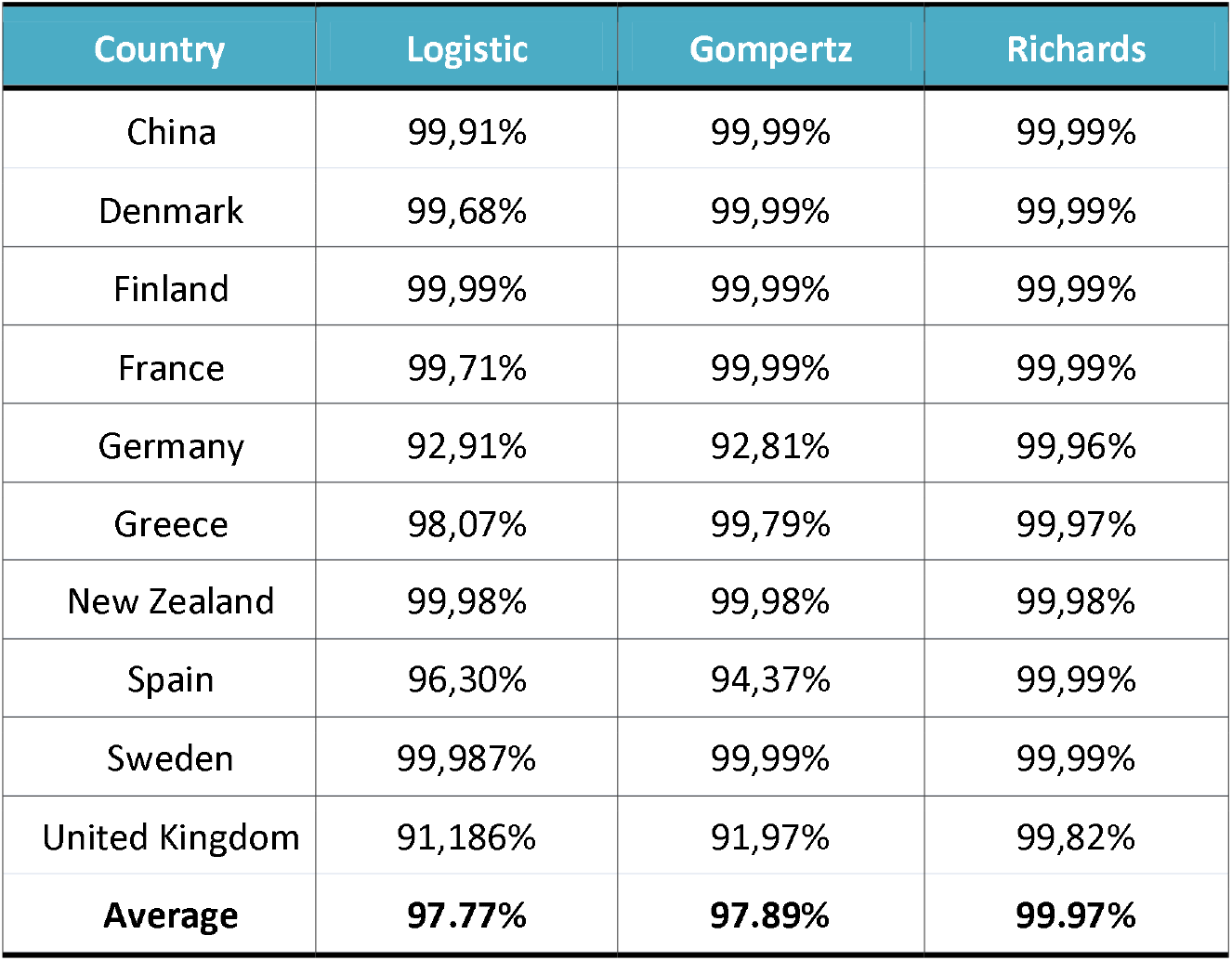
Descriptive Capacity of Growth Functions (R^**2**^)

The results in Table 3 allow us to answer the question - What is the best model? - from a descriptive point of view, with Richards model being the correct answer.

Figure 4. depicts the observational and theoretical data, both daily (left) and cumulative (right) in the 10 studied countries. Data is plotted for the overall interval of the first epidemic wave. Theoretical curves are displayed for the three studied models: Logistics, Gompertz and Richards.

**Figure 4.**
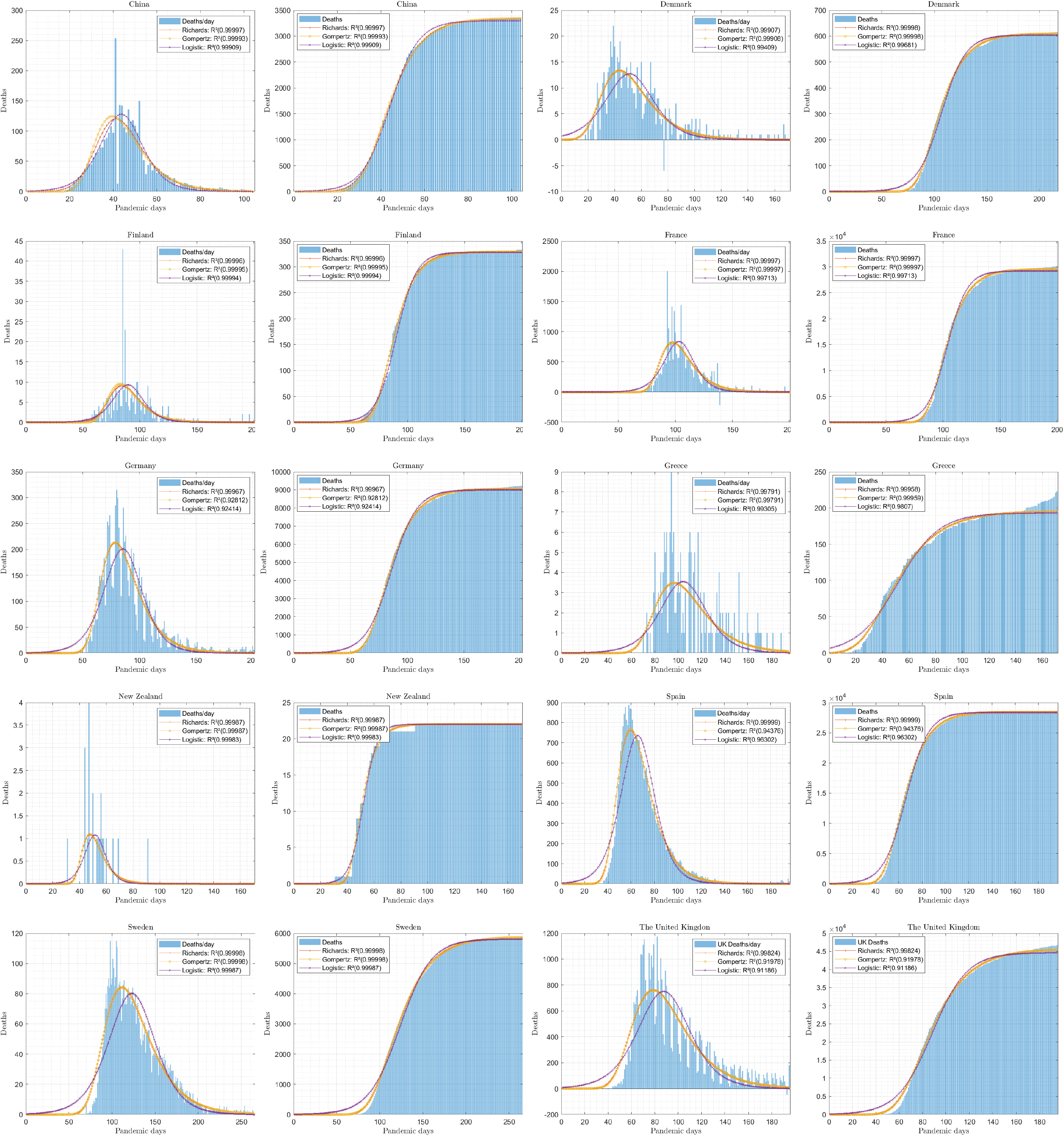
Descriptive capacity of growth functions: Observational (blue bars) and theoretical data (colored curves) for both daily (left) and cumulative (right) deaths in the 10 studied countries. Data is limited to the first epidemic wave.

The results shown in figure 4 allowed to perceive aspects that are not possible to see in the results shown in Table 3, such as:

- Richards and Gompertz models in most countries generate overlapping curves that are not possible to differentiate visually.
- The descriptive errors of the models are much better seen in the daily data than in the cumulative data, which should be taken into account in this type of study. In particular, the error of these models is greater in the initial and final phases of the epidemic wave, decreasing in the critical phase, and
- The logistic function suffers from a shift to the right of the peak of daily deaths in all cases, which is not the case with the other two models. This is probably due to the fact that the model is symmetric, while the data distribution is not symmetric in these countries.

Note, that the last two observations above are the answer to the stated question - What are the descriptive limitations of these models?

### Results of the Predictive Capacity of the Growth Models

In this section, we present results that allow us to answer the following questions: How do the studied models predict the total number of deaths in a complete epidemic wave in the selected countries? How far in advance do the models studied predict the total number of deaths in a complete epidemic wave in the selected countries, with a predefined tolerance for error? Which model is able to predict it earlier and more accurately? What are the predictive limitations of these models?

Table 4 shows the predictive metrics collected applying the best growth functions to each country.

**Table 4:** Results of the study of the forecasting capacity of growth functions.

| Country | Wave peak day ($d_{peak}$) | Day of prediction ($d_p$) | Wave ending day ($d_{end}$) | Anticipation days ($d_{ant}$) | Relative Anticipation ($R_{ant}$, %) |
| --- | --- | --- | --- | --- | --- |
| China | 42 | 65 | 80 | 15 | 39.5% |
| Denmark | 39 | 75 | 150 | 75 | 67.6% |
| Finland | 85 | 105 | 150 | 45 | 69.2% |
| France | 100 | 120 | 150 | 30 | 60.0% |
| Germany | 80 | 90 | 150 | 60 | 85.7% |
| Greece | 100 | 150 | 160 | 10 | 16.7% |
| New Zealand | 50 | 70 | 80 | 10 | 33.3% |
| Spain | 60 | 80 | 120 | 40 | 66.7% |
| Sweden | 100 | 130 | 200 | 70 | 70.0% |
| United Kingdom | 80 | 140 | 150 | 10 | 14.3% |

Summarizing, six of the ten countries were predicted with more than 60% of anticipation and the other four with less than 40%. The maximum achieved anticipation was 85.7% in Germany while the minimum anticipation was 14.3% in United Kingdom.

In figure 5 we show the evolution of the predicted total number of deaths, *D*_*pred*_(t), for the three optimized models in each country. For reference, the real death toll, *D*(=Target), was indicated with a horizontal red line, and the upper, (1 + *ε*/100)*D*, and lower, (1 *− ε*/100)D, limits of the error tolerance band with horizontal green lines.

**Figure 5.**
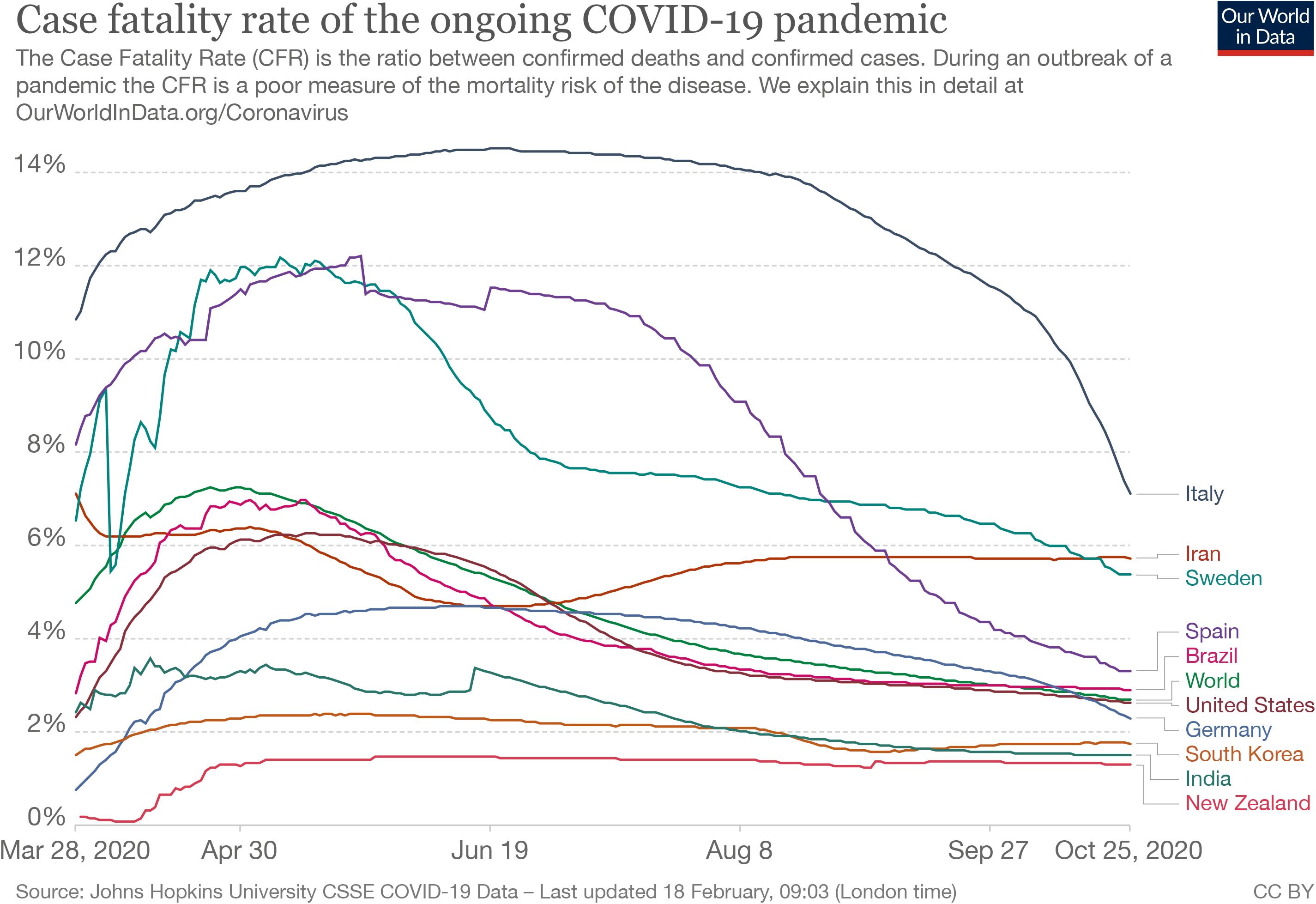
Convergent evolution of the predicted total number of deaths, *D*_*pred*_(t), for the three optimized models in each country.

Our first observation is that the predictions of the optimized models of Richards and Gompertz are practically indistinguishable in all countries in the final convergent phase. However, we prefer not to draw any conclusions due to the small number of countries considered in the study.

Second, as commented in the Materials and Methods section, at the beginning of the epidemic episode, the predicted value fluctuates from day to day until it starts to vary smoothly and converges monotonically to a value very close to the real number of deaths, *D*, in all cases. It should be remarked that in some countries (New Zealand, Greece, Finland, France), spurious fluctuations occurred also in the monotonically convergence period, mainly when using the logistic function (Greece, Finland, France). Fluctuations in New Zealand occurred with the Richards function.

Interestingly, in eight of the ten countries the final convergence occurs from the bottom for the three models, but in two countries (China and Finland) the convergence to the expected value of the Richards and Gompertz models is from the top.

Regarding the final predictive accuracy of the models, Table 5 shows the results obtained with the three growth model in the ten countries.

**Table 5:** Final Predictive Accuracy of Growth Functions (%)

| Country | Logistic | Gompertz | Richards |
| --- | --- | --- | --- |
| China | 98,27% | 99,94% | 99,94% |
| Denmark | 98,05% | 98,86% | 98,86% |
| Finland | 98,50% | 98,80% | 98,80% |
| France | 97,63% | 98,34% | 98,34% |
| Germany | 97,35% | 98,18% | 98,18% |
| Greece | 95,85% | 98,96% | 98,96% |
| New Zealand | 100.00% | 100.00% | 100.00% |
| Spain | 99,13% | 99,34% | 99,34% |
| Sweden | 98,81% | 99,98% | 99,98% |
| United Kingdom | 95,49% | 97,74% | 97,74% |
| Average | 97,91% | 99,02% | 99,02% |

In summary, the minimum final predictive precision is 95.49% corresponding to the logistic model with data from the United Kingdom, and the maximum is 100.00% for all models in New Zealand. According to the average final precision per model, Richards and Gompertz, again, have the same and best performances with an average final precision of 99.02%. These results confirm that the real-time prediction of the number of deaths is very stable in all models and in all countries and converges with very high accuracy to the real value.

Therefore, it is possible now to answer the question: Which model is able to predict earlier and more accurately? The results in the ten countries suggest that the Gompertz model has the best performance. Our conclusion is based on the fact that, even though it is very close to the Richards model in both relative anticipation and final accuracy, it has one less parameter and did not show spurious fluctuations in the convergent stage.

## Conclusions

The objective of this work was to investigate, on a statistical basis, the ability of classical growth functions to model the data from the COVID-19 pandemic. At the time of the survey (late October 2020), the pandemic was still ongoing and more than a wave of cases and deaths occurred in several countries.

The 3 factors that motivated this research are: (1) The high number of articles published with this type of functions, (2) The lack of a classification of the different types of problems that can be faced with data analysis techniques in this specific context, and (3) The lack of a methodology based on quantitative metrics to measure performance in solving these different types of problems.

To establish a classification of the problems, it was necessary to formalize the concept of epidemic wave portrayed in the daily data, establishing its basic characteristics. In particular, the division into two phases, initial-increasing and final-decreasing, and the characterization of the phases according to their intensity in low and high.

Once this definition was made by mining data from all available countries, it was possible to establish 4 types of waves and verify the independence between the two phases of the epidemic waves.

Thus, it was possible to determine the resolvability of different types of problems depending on the stage of the epidemic wave, the biggest conclusion being the impossibility of solving the long-term forecasting problems (the total values of an epidemic wave) with data from the first phase only. This, in fact, implies that it is not worth investing efforts to solve this type of indeterminate problem, at the moment that puts in doubt the capacity of generalization of all published methods that managed to make this type of prediction satisfactorily using only data before the peak of the wave. Data analysts are well aware that data cannot tell us what they don’t know, but that under torture, they are able to answer what we want to hear.

Once the theoretical and methodological support is established, we focus on evaluating, using the metrics specifically designed for these types of problems, the performance of 3 classic growth functions: Logistics, Gompertz and Richards (the latter treated as a generalization of the previous two) for describe the path of the epidemic (problem type 1) and to make long-term predictions (problem type 5) during the second phase of the epidemic waves. We used data from 10 countries, 6 of them with more than X daily cases on the peak day.

The results show a generalized underperformance of the logistic function in all aspects and places the Gompertz function as the best cost-benefit alternative, as it has performance comparable to the Richards function, but it has one less parameter to be adjusted, in the process of regression of the model to the observed data.

However, as the basic rules of data analysis recommend, before choosing a model for a predictive problem, evaluate the descriptive capacity (problem type 1) of several models and then choose the one with the highest descriptive capacity to solve your predictive problem.

## Data Availability

we used only public data available on the WHO website

## Acknowledgments

We wish to confirm that there are no known conflicts of interest associated with this publication and there has been no significant financial support for this work that could have influenced its outcome. VF is supported by CAPES (88882.349290/2019-01) and a Flagship grant from the South African Medical Research Council (MRC-RFA-UFSP-01-2013/UKZN-HIVEPI).

